# Contextual factors and implementation strategies for a biomarker-augmented alcohol screening with brief intervention and referral to treatment (SBIRT) program for HIV-affected adolescents in Zambia: a qualitative study guided by RE-AIM / PRISM

**DOI:** 10.1101/2025.08.07.25333191

**Authors:** Alejandra Paniagua-Avila, Tukiya Kanguya, Chanda Mwamba, Judith A. Hahn, Carl Latkin, Geetanjali Chander, Silvia S. Martins, Saphira Munthali, Michael G McDonell, Anjali Sharma, Jeremy Kane

## Abstract

**Introduction:** Screening, Brief Interventions and Referral to Treatment (SBIRT) programs reduce unhealthy alcohol use among adolescents. However, self-report screening alone may lead to false negatives and low service use, especially in HIV care settings. This study explored the contextual implementation factors and strategies of an alcohol biomarker-augmented SBIRT program for HIV-affected adolescents in Zambia, where alcohol use and HIV prevalence are high.

**Methods:** We conducted key informant interviews (n=7) with mental health providers and policymakers and focus groups (n=16 groups; 10-11 participants each) with healthcare providers, adolescents, and caregivers, guided by a case vignette of the biomarker-augmented SBIRT program. Thematic analysis followed the implementation frameworks Reach, Effectiveness, Adoption, Implementation, Maintenance (RE-AIM) and Practical, Robust Implementation and Sustainability Model (PRISM).

**Results:** Participants perceived the SBIRT program as appropriate for adolescent alcohol use. Key contextual factors included: lack of alcohol treatment programs, community stigma against HIV and alcohol use, and robust implementation infrastructure through HIV healthcare. Strategies to enhance acceptability included making alcohol screening universal to avoid labeling adolescents, privacy and confidentiality during biomarker sampling, and peer-led age-matched counseling at screening. To enhance reach, participants suggested designing the program with attention to gender-specific needs and integrating it into HIV healthcare and alcohol use hotspots (e.g. schools).

**Conclusions:** Implementation strategies should be designed to reduce stigma, build trust, engage adolescents across genders, and reach youth through clinical and community channels. Future research should define how to select, train, and evaluate peer counselors and assess the effectiveness of alcohol biomarkers within SBIRT programs in motivating behavior change.

## 1. Introduction

Many adolescents living in Southern Africa experience the double burden of HIV infections and substance use [1, 2]. While the incidence of HIV has declined, the region is still home to 85% of adolescents living with HIV worldwide [3, 4]. Addressing the behavioral determinants of HIV infection, such as unhealthy alcohol use, is imperative to control the HIV epidemic among adolescents in Southern Africa [5]. Studies have shown that the prevalence of alcohol use among people living with HIV is high. Meta-analyses have estimated that the average prevalence of alcohol use disorder among people living with HIV is 29.80% (95% confidence interval, CI: 24.10-35.76) globally and 22.03% (95% CI: 17.18-28.67) in Africa [6, 7]. In addition, meta-analyses have suggested that behavioral interventions targeting alcohol use among people living with HIV may help to reduce alcohol use and improve HIV-related outcomes, such as medication adherence and viral load [8]. Together, these studies show that alcohol use is a highly prevalent behavioral determinant of HIV harmful outcomes that could be intervened upon to improve health. However, alcohol use among people living with HIV, and particularly adolescents in Southern Africa, has received limited public health attention [9].

Adolescents experience a high burden of unhealthy alcohol use globally [10]. This burden is expected to continue increasing, particularly in low- and middle-income countries (LMICs) including the Southern African region [9, 11]. People who use alcohol or substances are disproportionally affected by risky sexual behaviors, sexually transmitted diseases, HIV transmission, and higher viral HIV loads [12, 13]. Moreover, alcohol use during adolescence increases the risk of developing alcohol use disorder later in life [14, 15]. Overall, unhealthy alcohol use can lead to societal costs related to healthcare expenses, lost productivity, and years of life lost globally [16]. Identifying and addressing alcohol use among adolescents may prevent the later development of alcohol use disorder, HIV transmission and higher viral loads, and other comorbid conditions later in life. However, few adolescents in Southern Africa or other LMICs receive services for alcohol use, with a treatment gap of up to 90% [9].

One response to the increasing alcohol use burden is early identification and intervention. Screening, Brief Intervention and Referral to Treatment (SBIRT) programs are an integrated public health approach to the delivery of universal screening and early intervention and treatment for alcohol use [17] that may be integrated into HIV care. SBIRT programs have been shown to be effective at reducing alcohol use among adults and adolescents [17–21]. SBIRT conceptualizes alcohol use as a continuum, ranging from risky alcohol use to alcohol use disorder [21, 22]. As such, SBIRT targets all types of unhealthy alcohol use, tailoring treatment intensity according to severity [21]. SBIRT consists of three steps: first, the individual is screened (S) for unhealthy alcohol use, commonly through validated self-reported questionnaires; second, if screening is positive, the individual receives a brief intervention (BI), consisting of counseling techniques aiming at motivating behavioral change; finally, depending on severity, the individual may be referred to treatment (RT) at a higher level of care [17, 21]. Despite limited data among adolescents, studies suggest that SBIRT programs may be effective at addressing unhealthy alcohol use in Southern Africa [23–25]. For example, a recent study in Zambia demonstrated that an SBIRT program featuring cognitive behavioral therapy delivered by lay counselors was effective at reducing unhealthy alcohol use among adults living with HIV within Lusaka, Zambia HIV clinics [23, 24]. However, SBIRT programs are yet to be integrated into routine HIV care services for adolescents in Southern Africa, missing the opportunity to prevent the development of alcohol use disorders and additional HIV-related negative outcomes.

Our team is preparing to deploy a new alcohol SBIRT program for HIV-affected adolescents integrated within HIV care services in Lusaka, Zambia. HIV-affected adolescents comprise adolescents living with HIV, adolescents who were orphaned by HIV, adolescents who live with someone who is HIV-infected, and adolescents who are at increased risk for HIV infection [26]. In partnership with the Center for Infectious Disease Research in Zambia (CIDRZ), we have chosen to focus exclusively on alcohol use, given the pilot nature of this study. During screening (S), we will conduct point-of-care rapid testing with urine-based ethyl glucuronide (EtG), an alcohol biomarker that provides qualitative (i.e., positive/negative) evidence of any recent (2-5 day) alcohol consumption within five minutes [27–29] along with self-report.

Utilizing biomarkers in conjunction with self-report may help to address common issues of self-reported alcohol screening approaches such as social desirability and recall bias, while allowing healthcare providers to establish rapport, facilitating the subsequent phases of the SBIRT program, including counseling and referral [30, 31]. Moreover, it may allow us to evaluate and compare the accuracy of self-report against biomarker-based alcohol screening as part of the larger study. During the brief intervention (BI), a counselor, such as a nurse or a physician, will provide personalized feedback and brief counseling aimed at reducing alcohol use, based on the test result and self-report [19]. The counselor may also refer the adolescent to more intensive treatment options, such as cognitive behavioral therapy, during the referral to treatment phase (RT) [17, 19]. We plan to pilot and integrate the alcohol SBIRT program into existing HIV care services for adolescents, in partnership with CIDRZ.

Although a biomarker-augmented SBIRT program has the potential to improve the accuracy of alcohol screening, to our knowledge the strategies to implement this program within HIV care services for adolescents in LMICs have not been explored. Thus, we sought to identify the multi-level factors that may influence the implementation of such a program for adolescents in Lusaka, Zambia, and elicit implementation strategies to enhance its acceptability, adoptability, and reach from adolescents, caregivers, healthcare workers, and policymakers.

### 1.1. Study setting

This study was conducted in Lusaka District, a densely populated urban center in Zambia. Adolescents make up 25.0% of the population in Zambia [32]. Lusaka, Zambia is characterized by high HIV prevalence among adolescents and young adults, combined with increasing numbers of adolescents who use alcohol or other substances [32]. A recent analysis of nationally representative data from Zambia found that among young adults living with HIV, the prevalence of unhealthy alcohol use was 21.6% and it was associated with reduced odds of viral suppression [33]. This study was co-led by CIDRZ, a research center that implements primary care- and community-based HIV programming in Lusaka. One such program, Youth Friendly Spaces, is a peer educator-led HIV intervention focused on providing HIV screening and prevention interventions to adolescents and young adults.

## 2. Materials and methods

### 2.1. Ethics Statement

This study was approved by the Columbia University Irving Medical Center (CUIMC) Institutional Review Board (IRB-AAAT5996) and the University of Zambia Biomedical Research Ethics Committee (REF-NO. 1814-2021). All participants provided verbal informed consent at the time of recruitment. Given that the study was low risk, in the case of adolescents aged 16 and 17 years, we obtained a waiver of caregiver / parental consent to protect adolescents’ confidentiality and to facilitate participation of adolescents with personal experiences with alcohol or substance. Researchers made sure that the participants freely participated, were not forced into the study, and collected data in private settings. Each participant received K100 (USD 3.80) and snacks for participating in in-depth interviews or focus group discussions. Participants were recruited from 24/NOV/2021 to 10/MAR/2022.

### 2.2. Overview of study design

In preparation for piloting of a biomarker-augmented alcohol SBIRT program, this study collected in-depth interviews and focus group discussions with a wide range of participants (adolescents, caregivers, service providers, and policymakers) to explore knowledge, attitudes, and behaviors around alcohol and substance use among adolescents, as well as existing policies, services, and programs for addressing alcohol and substance use by adolescents in Zambia. In this paper, we present findings from a secondary analysis with a subset of qualitative data to better understand the multi-level contextual implementation factors and implementation strategies relevant to a biomarker-based alcohol SBIRT program for adolescents. Our secondary analysis was guided by the Practical, Robust, Implementation and Sustainability Model (PRISM) and the Reach, Effectiveness, Adoption, Implementation, Maintenance implementation science frameworks (RE-AIM/PRISM). **Study Box 1** summarizes the PRISM dimensions included in this study, their definition, and their operationalization through selected examples of interview or focus group questions. The Consolidated criteria for reporting qualitative research (COREQ) [34] is included as **Supplementary material 1.**

**Study Box 1.** RE-AIM and PRISM dimensions, definition, and examples of interview or focus group questions.

| Domain | What we are assessing |
| --- | --- |
| **PRISM: Multi-level contextual factors** |  |
| Patient, caregivers, and community characteristics | - Current substance use practices<br>- Current help seeking behavior<br>- Stigma |
| Patient, caregivers, and community perspectives | - Perceived need for the program<br>- Perspectives of the program |
| Organizational characteristics | - Identification, referral, and care for adolescents who use alcohol or substances |
| Organizational perspectives | - Perceived need for the program |
| Implementation and sustainability infrastructure | - HIV services for adolescents<br>- Preventative services<br>- Treatment services |
| External environment (policy, resources, guidelines, incentives) | - National legislation, policies and public funding for alcohol and substance use |
| **RE-AIM: Strategies and recommendations** |  |
| Implementation strategies for enhancing selected implementation outcomes | - Strategies for enhancing program <u>reach</u><br>- Strategies for enhancing program <u>adoption</u> and <u>acceptability</u> |

### 2.3. Implementation science frameworks

Our analysis was guided by two implementation science frameworks: the Practical, Robust, Implementation and Sustainability Model and the Reach, Effectiveness, Adoption, Implementation, Maintenance (RE-AIM/PRISM). Implementation scientists frequently utilize RE-AIM to assess key implementation outcomes of evidence-based interventions (Glasgow 2019). PRISM, a contextual expansion of RE-AIM, was developed to evaluate multi-level contextual factors relevant to the implementation of complex interventions (Feldstein 2008, Rabin 2022, Glasgow 2019). Concepts from implementation research inform PRISM, the diffusion of innovations theory, models of chronic care, and quality improvement (Rabin 2022). RE-AIM is an evaluation framework, while PRISM may be classified as an evaluation and determinant framework (Nilsen 2015). Here, we utilized PRISM to describe the following dimensions or determinants of implementation: community characteristics and community-level perceptions of the SBIRT program, including those from adolescents and caregivers; organizational characteristics and organizational-level perceptions of the program, including those from policymakers, professional healthcare workers and community healthcare workers; implementation and sustainability infrastructure; and external environment. Additionally, we used RE-AIM to categorize participant-identified implementation strategies by the implementation outcome that they addressed. We focused on the following RE-AIM implementation outcomes: Adoption (Acceptability, Adoptability) and Reach (See **Study Box 1**).

### 2.4. Participants

Through purposeful sampling, we recruited participants representing a wide range of individuals knowledgeable about the context where alcohol use and HIV treatment services take place in Lusaka, Zambia. Participants included seven key informants and 161 focus group discussions. Table 1 summarizes the characteristics of study participants. Key informants were mental health experts (n=4) who led nonprofit mental health and substance use programs, and policy-makers (n=3) affiliated with public health institutions. Focus group participants were HIV-affected adolescents (ages 16-24 years), caregivers of adolescents, professional healthcare workers involved in HIV care or programming for adolescents, peer educators, community counselors, mental health professionals, and policymakers knowledgeable about alcohol and substance use among adolescents in Lusaka, Zambia. Adolescents were considered HIV-affected if a person was receiving home-based HIV care in their immediate family and if they were accessing HIV prevention and treatment services at the study clinics. Potential adolescent and caregiver participants were identified by CIDRZ research and clinical staff in charge of home-based visits and Adolescent Friendly Spaces, who were then referred to research assistants. CIDRZ research assistants identified healthcare workers, counselors, and policymakers. After identifying potential participants and prior to initiating study procedures, CIDRZ research assistants performed verbal informed consent. All study participants included in this study provided verbal informed consent.

**Table 1.** Characteristics of study participants.

| Overview | In-depth interviews (n=7) |  | Focus group discussions<br>(n=16; 10-11 participants each) |  |  |  |
| --- | --- | --- | --- | --- | --- | --- |
|  | MH experts | Policy-makers | Adolescents<br>(n=4) | Caregivers<br>(n=4) | Professional healthcare workers<br>(n=4) | Peer educators and community counselors<br>(n=4) |
| n=168 | 3 | 4 | 40 | 40 | 41 | 40 |
| Female, 105<br>(62.5%) | 1 (0.33%) | 1 (25.0%) | 13 (32.5%) | 32 (80.0%) | 34 (82.9%) | 24 (60.0%) |

### 2.5. Qualitative data collection

The CIDRZ study coordinator (TK), a global health researcher who was trained and experienced in qualitative methods, collected data from October 2021 to February 2022. We first conducted in-person in depth interviews with key informants who represented mental health professionals and policymakers. Interviews lasted up to 2 hours. The study coordinator conducted the in-depth interviews in English, the official language for education and business in Zambia, within private locations selected by the mental health experts and policymakers.

We then performed in-person focus group discussions with separate groups of adolescents (ages 16-24), professional healthcare workers, peer educators, and lay counselors. Focus Group Discussions lasted up to 3 hours. Groups with professional healthcare workers were conducted in English, while other groups used a mix of English, Nyanja, and Bemba. This multilingual switching is common in Lusaka’s social settings, where people often shift languages to convey their thoughts effectively and comfortably. Research Assistants were fluent in both English and local languages (Bemba and Nyanja) and ensured privacy and comfort for participants, facilitating open and honest discussions. The choice of private rooms within the health facilities offered a familiar and secure setting to participants, encouraging their active engagement during the sessions. Interviews and focus group discussions were audio recorded and then transcribed verbatim by CIDRZ research assistants.

In-depth interviews and focus group discussions elicited information about three areas related to adolescent alcohol and substance use: 1) at-risk groups, common practices, consequences, and community responses; 2) existing community- and health system-based services, and policies; and 3) perceptions and recommended strategies for the implementation of a potential biomarker-based alcohol SBIRT program for adolescents. To describe the SBIRT program in the third section, we used a case vignette developed by the study coordinator (TK) and research assistants (See **Study Box 2**). This paper focuses on themes related to the implementation of the SBIRT program guided by the RE-AIM/PRISM frameworks, elicited primarily in the third section of the interviews and focus group discussions, and through selected questions in the first and second sections. Broader results about the remaining information will be presented in a separate paper.

##### Study Box 2. Case vignette describing the biomarker-augmented SBIRT program integrated into HIV services

I would now like to ask for your input on a potential program that could be used to ask about alcohol use and test for alcohol use among adolescents during regular HIV care services that they receive. Sometimes adolescents do not feel comfortable talking about alcohol that they might consume. This program would ask adolescents about alcohol that they might use and will also use a urine-based test called EtG that can provide some indication about whether the adolescent has taken any alcohol in the past few days. This is not a program that currently exists but one that could possibly exist in the future, and we would like to get your opinions on this. Now I will describe an example of what this program might look like and show you some pictures as an example.

I would like you to imagine an adolescent named Francis. Francis is 16 years old and is living with HIV. He receives care at Kanyama clinic. During a regular visit to Kanyama clinic to receive his medication and meet with a nurse, he is asked about whether he consumes alcohol. The nurse asks him questions about whether or not he has taken alcohol recently, how often he drinks alcohol, and how much he drinks on occasions when he takes alcohol.

After answering these questions, the nurse asks Francis to use a private room and to provide a urine sample in a specimen container like this. After providing the urine sample, the nurse will place the tip of an EtG dipstick that looks like this into the urine sample container for about 15 seconds and then place it on a flat surface like this.

Over the next five minutes, the nurse may ask Francis about other things, or they may take care of other business for the appointment, such as collecting refills on his medication from the pharmacy. After five minutes, the nurse will review the dipstick results (show picture). If the test is positive, this means the adolescent may have drunk alcohol in the past few days. If the test is negative, this means that the adolescent probably did not drink any alcohol in the past few days.

The nurse sits down with Francis and reviews the results of the EtG test with him and what they mean (show picture). She may also discuss with Francis if the results of the EtG test are different from his responses to the questions he gave on alcohol use. For example, if Francis responded to her earlier questions that he never takes alcohol, but the EtG test is positive, she may ask him if maybe he had taken alcohol but felt uncomfortable talking about it or maybe if he forgot he had taken alcohol. If Francis has consumed alcohol, the nurse may provide him a brief intervention in which they discuss why he drinks and some strategies for reducing the amount and how often he drinks. She may also connect him to additional services if he drinks a lot.

### 2.6. Qualitative data analysis

Three study team members (APA, TK, CM), experienced in qualitative methods, performed a thematic inductive-deductive analysis of interview and focus group discussion data. The inductive coding phase focused on identifying emerging ideas from qualitative transcripts through open coding. Each analyst independently read one interview and one focus group transcript to identify and annotate emerging words, phrases, and ideas. The analysis team met multiple times to review and discuss the main ideas and group them into codes. Then, two analysts tested the codes in a subset of transcripts (APA, TK), solving discrepancies with the help of a third analyst (CM), and further refining the codes until we did not identify new themes, suggesting that we reached saturation. The final codebook (See **Supplementary material 2)** was applied to all transcripts using NVivo Software.

We then performed a sub-analysis to identify multi-level contextual factors relevant to the SBIRT program’s implementation and recommendations for the program’s implementation. We followed a deductive approach in which a subset of previously identified inductive themes related to the RE-AIM / PRISM dimensions were re-analyzed following a thematic analysis. All in-depth interviews and focus group discussion transcripts were included in this sub-analysis.

## 3. Results

### 3.1. Participants

Participants included seven key informants and 161 focus group discussants. **Table 1** summarizes the characteristics of study participants. Key informants were mental health experts (n=4) who led nonprofit mental health and substance use programs, and policymakers (n=3) affiliated with public health institutions. Focus group discussants were adolescents (n=40), adolescent caregivers (n=40), professional healthcare workers (n=41), and peer educators and community counselors (n=40), including individuals who could conceivably provide alcohol screening and intervention services within HIV care. Most key informants were male (n=5; 71.4%), while most focus group participants were female (n=103; 64.0%). Most adolescent focus group participants were male (n=27; 67.5%).

### 3.2. Multi-level implementation factors

The following sections present key results related to multi-level factors relevant to the implementation of an alcohol SBIRT program for adolescents in Lusaka, Zambia, organized by dimensions of the PRISM framework (**See Table 2**).

**Table 2.** Multi-level implementation factors of alcohol screening intervention.

| PRISM dimension | Exemplary quotes for themes within PRISM domains (Data source, type of participant) |
| --- | --- |
| Adolescent, caregiver, and community characteristics | <u>Alcohol and substance use practices among adolescents:</u><br>*“So, when struggling with a trauma maybe abuse, physical abuse, sexual abuse... to cope they will find the drug substance which they will take to calm them down”* (Quote 1 - KII, Mental health professional)<br>*“How can he stop them from taking alcohol when there is nothing to do and so they take alcohol?”* (Quote 02 - FGD, Caregiver)<br>*“Poverty is one of them and influence from peers ... I think I will be a laughingstock if I am not participating in what my friends are doing and there’s nothing literally that I am doing and it’s just better that I participate. So, I think influence from friends plays a role”* (Quote 03 - FGD, Adolescent)<br><u>Support seeking:</u><br>*“These things are just harming our lives. It was a mistake. All this was a mistake and at the time we thought this was the way to go but it was a mistake and at this point we now are, we cannot turn back the hands of time because it’s too late”* (Quote 04 - FGD, Adolescent)<br>*“Yeah, so if I find out that they are abusing drugs, I’ll try to keep them busy, keep their mind away from drugs and just having been entertained and let them get the proper the proper knowledge they need and all that so they should learn in the understand the effects of drugs”* (Quote 05 – FGD, Adolescent)<br><u>Stigma:</u><br>*“I think the better term is that they are ‘reject’s in the society’ because no one wants to be near them. We are scared because we do not know what they will do to you next”* (Quote 06 - FGD, Professional healthcare worker)<br>*“I am going to refer to the child that abuses Bostick whom I made reference to earlier. I spoke to the mother to bring the boy to Chainama Hospital for further management but knowing that the hospital deals with mental patients she decided to abandon the idea for fear of being stigmatized in the community that she has a mad son”.*<br>(Quote 07 - FGD, Community healthcare worker) |
| Adolescent and caregiver perspectives of the intervention | <u>Perceived need for the intervention:</u><br>*“I think it’s okay and it would help him to keep track and it will help the caregiver to know how often “Francis” has reduced, or is “Francis” still drinking, taking alcohol at regular basis; of which, in as much as the care giver would want to know that, but it will somehow help “Francis” overcome beer challenges. Why? Because the first time it will test positive maybe, then the second time, okay, he will have that feeling to say ‘every time I go there, they have to test for alcohol,’ so you find that even the intake probably it might reduce - this as my assumption, but I think it’s a good thing”* (Quote 08 - FGD, Adolescent)<br>*“I would encourage you that soon it should start. Those that are living with HIV are just a small number and those without HIV are a bigger number, so you should do it for everyone”* (Quote 09 - FGD, Caregiver)<br><u>Concerns:</u> |
|  | *"It will be uncomfortable in the sense whereby I have only gone there for a specific thing and then that person has started raising other issues. So in fear of being looked down on, he will say 'No, I do not take alcohol'" (Quote 10 - FGD, Adolescent)* |
| Organizational characteristics | <u>Identification, referral, and care for adolescents who use alcohol or substances:</u><br>*"To say the truth not many people, know about these programs because there is lack of information and we generally not created awareness in the communities about these programs. So, it's like we have abandoned these young people since we are not doing much to provide the information as providers" (Quote 11 - FGD, Community healthcare worker)* |
| Organizational perspectives of the intervention | <u>Perceived need for the intervention:</u><br>*"I have hope in your desire to implement this program. It should not just end with this focus group discussion. It should be implemented because we have a lot of adolescents who are abusing alcohol and need help" (Quote 12 - FGD, professional healthcare worker)* |
| Implementation and sustainability infrastructure | <u>Preventative services:</u><br>*"Women have a lot of activities to participate in, for instance, they are taught how to make bar soap. Then after that they sell, then they get commissioned from there. We also have DREAMS program for female adolescents, where they are taught how to make earrings, hats, and shoes, and handbags. So for men and with your research think about 'How are we going to help male adolescents?' because women have a lot of ways when it comes to helping them. (Quote 13 - FGD, community healthcare worker)*<br><u>Treatment services:</u><br>*"The ones that are in charge for curbing drugs, I would say they are putting more effort but for me they are failing. So, I would say that we need more programs, because it is not just HIV, there is a lot happening in these communities, reason why we need more programs, maybe to involve more people" (Quote 14 - FGD, community health worker)*<br><u>HIV services for adolescents:</u><br>*"For adolescents we have youth friendly space, so this building is open for young people we mostly talk about drug abuse, HIV and STIs and other issues affecting young people" (Quote 15 – FGD, Community healthcare worker)* |
| External environment | <u>Legislation, policies, and funding:</u><br>*"So, as one of the measures to cut down or reduce accessibility [to alcohol] is pricing so we have been calling government on a number of platforms to say: 'Can you increase the price by increasing the tax bracket for alcohol?'" (Quote 16 - KII; Mental health expert)*<br>*"So, the type of laws we have can be a hindrance, in the sense that some people fail to access the service because they will think probably, I will get information and report them to Drug Enforcement Commission (DEC). So, there are certain drugs that need to be ... There are two words I want to use ... There's a difference between the legalization and decriminalization" (Quote 17 - KII; Mental health expert)* |
| FGD – Focus Group Discussion<br>KII – Key informant interview |  |

#### 3.2.1. Adolescent, caregiver, and community characteristics

##### Alcohol and substance use practices among adolescents

Adolescents expressed using alcohol or other substances, to feel ‘pleasure’, ‘high’, or have ‘new experiences’ in groups. Girls frequently used alcohol during sex as they believed it enhanced boys’ satisfaction. In addition, some adolescents mentioned that alcohol use may lead to risky sexual behaviors such as having multiple sexual partners. Adolescents also expressed using alcohol or other substances to ‘cope’ or ‘escape’ from difficult experiences such as failing a school test or past traumatic experiences, such as losing a close relative or experiencing sexual abuse (see **Table 2, Quote 01**). Key informants, healthcare workers, and community counselors identified poverty, not being enrolled in school, unemployment, and perceived lack of parental care or love as structural factors that put adolescents at risk for using alcohol or substances. Being diagnosed with HIV was frequently identified as a precursor, along with peer pressure and school bullying, to start using alcohol or substances. Caregivers considered that the lack of recreational activities for adolescents is one of the facilitators for alcohol and substance use (See **Table 2, Quote 02).** In turn, substance use led to risky sexual behaviors, such as not using condoms or having multiple sexual partners, which seemed to increase the risk of transmitting or acquiring HIV. Participants agreed on the main sources of alcohol or substances for adolescents, which included schools, illegal bars, and other peers. Adolescents preferred using low-cost alcohol or substances that could be easily obtained. Adolescents expressed a strong preference for consuming alcohol or substances in private spaces, such as vacant houses, below bridges, parks, and behind bushes or taverns. Schools were also a source and a hot spot for consuming alcohol or substances during recess and school time (see **Table 2, Quote 03**).

##### Support-seeking practices

Most adolescents who reported using alcohol or substances expressed a strong desire to stop but found it difficult to do so, and believed it was extremely challenging to reduce using them after having initiated (See **Table 2, Quote 04**). Caregivers of adolescents who used alcohol or substances expressed their concerns and shared their failed attempts to help their children. Most adolescents and caregivers did not know how to seek help through existing programs. Those who knew how to do so expressed difficulties accessing programs due to geographic or economic limitations. However, many adolescents said that if they had a peer with alcohol or substance use problems, they would try to support them by keeping them busy, listening, chatting, or joking with them, inviting them to play a sport, or inviting them to community-based programs such as Youth Friendly Spaces (see **Table 2, Quote 05**).

##### Stigma against adolescents who use alcohol or substances

Adolescents with alcohol or substance use problems were seen as ‘outcasts’, ‘rejects’, or ‘youngsters with no future’ by community members and healthcare workers (See **Table 2, Quote 06**). They were often considered ‘problematic’ adolescents who often insulted community members or stole to support their substance or alcohol use. Community health workers indicated that community members often withdrew from adolescents who used alcohol or drugs and avoided interacting with or being associated with them. Community and professional healthcare workers frequently identified stigma as a major challenge to accessing services for adolescents who use alcohol or substances (see **Table 2, Quote 07**).

#### 3.2.2 Adolescent and caregiver perspectives of the SBIRT program

##### Perceived need for a screening program

Overall, there was consensus among adolescents and caregivers about the urgent need for an alcohol SBIRT program. From the perspective of caregivers, adolescents in Lusaka, Zambia were at risk for alcohol use disorder. A screening program would help to identify adolescents that are currently unidentified. Caregivers considered that the screening should be made mandatory for all adolescents living with HIV and those without HIV, to avoid labeling and stigmatizing those who use alcohol and those who live with HIV. Adolescents thought that the program could help HIV-affected adolescents reduce drinking, which often makes them forget their HIV follow-up appointments (See **Table 2, Quote 08**). Caregivers thought that the program should go beyond HIV-affected adolescents to reach all adolescents in need (See **Table 2, Quote 09**)

##### Adolescent concerns about the screening program

Adolescents’ perspectives on an alcohol screening program ranged from those who accepted screening as routine, comparing it to screening tests for sexually transmitted infections (STIs), to adolescents expressing concerns about program characteristics or potential for unintended consequences. For example, some participants thought that adolescents could feel embarrassed or offended about providing urine samples or sharing information about their alcohol use. If healthcare workers were rude or lacked compassion when asking for urine samples, adolescents who use alcohol could feel judged and misunderstood. In response, some adolescents could stop using alcohol in preparation for their HIV routine appointments, which could affect the screening accuracy. Finally, some adolescents expressed concerns about the potential impact of the program on ART, such as delaying antiretroviral therapy (ART) initiation or reducing the frequency of ART use due to adolescents’ fear of being identified as alcohol users (see **Table 2, Quote 10**).

#### 3.2.3 Organizational characteristics

##### Identification, referral, and care for adolescents who use alcohol or substances

Overall, there was consensus among participants that there is limited availability of services and programs for adolescents who use alcohol or substances. Available programs were focused broadly on primary prevention or specifically on care for adolescents with established alcohol or substance use problems or overdoses, overlooking early identification and treatment for adolescents without an overt substance disorder. All types of healthcare workers indicated that most programs focused on alcohol or substance use have failed. For this reason, most adolescents who need care are not identified and do not receive care. Moreover, healthcare workers indicated that most adolescents and their caregivers did not seek care due to a lack of awareness about available services, long distances from the community to hospitals, and stigma against seeking mental health and alcohol or substance use programs (see **Table 2, Quote 11**).

#### 3.2.4 Organizational perspectives of the intervention

##### Perceived need for the intervention

Policy makers and mental health, professional, and community healthcare workers agreed on the urgent need for implementing programs to identify and treat adolescents with alcohol or substance use problems (See **Table 2, Quote 12)**. Participants thought that integrating an alcohol SBIRT program into HIV care could help adolescents reduce alcohol use, which would increase service engagement, reduce the risk of missing their HIV-related appointments, and improve ART use. Community healthcare workers thought that the screening intervention on its own could help adolescents reduce their alcohol use, as they would prepare for their appointments by reducing alcohol drinking.

#### 3.2.5 Implementation and sustainability infrastructure

##### Preventative services

Participants described a series of preventative services or programs that aimed to reduce alcohol or substance use initiation among adolescents in Zambia led by community nonprofit organizations or civil associations. Preventative services commonly took place in churches or community settings within urban compounds. Community healthcare workers indicated that services often had a broad scope and focused on other health outcomes in addition to alcohol or substance use, such as teen pregnancy and HIV prevention. Specific activities included support groups that provided education about the ‘dangers of alcohol and early pregnancy’ or emotional support for adolescents living with HIV. Preventative services may also focus on ‘keeping adolescents busy’ through sports, physical activity, dancing, or educational conversations about avoiding substance or alcohol use. Since most programs were designed to target girls and female adolescents, many healthcare workers and adolescents expressed the need for additional preventative programs targeting boys and male adolescents (see **Table 2, Quote 13**).

##### Treatment services

Participants identified a few services tailored to adolescents who were already using alcohol or substances, led by religious organizations, and held within large public hospitals or inpatient rehabilitation facilities. Some treatment services also provided counseling and educational sessions to adolescents as well as interventions following ‘harm reduction’ principles. However, as previously indicated, most healthcare providers reported that most past and existing programs have failed in part due to stigma and geographic barriers, pointing at the need for a different approach to programs to address alcohol use and substance use (**See Table 2, Quote 14).**

##### HIV services for adolescents

Mental health, professional and community healthcare workers agreed on the need to deliver alcohol and substance use programs within existing community- or primary care-based HIV services for adolescents. For example, a SBIRT program could be integrated into Youth Friendly Spaces. Specifically, a SBIRT program could be integrated into the educational and counseling activities aimed at preventing substance use among adolescents affected by HIV, or into the workflow of HIV care and ART delivery (see **Table 2, Quote 15**).

#### 3.2.6 External environment

##### National legislation, policies and public funding for alcohol and substance use

Mental health experts and policymakers described multiple barriers to developing and funding alcohol and mental health programs for adolescents. First, there was a lack of national legislations and policies supporting the population that uses alcohol or drugs. For this reason, there was a lack of funding allocated to alcohol and mental health services. Second, access to alcohol among adolescents was facilitated by it being inexpensive, due to the lack of alcohol taxes (See **Table 2, Quote 16**). Finally, substance use is criminalized in Zambia, which often discourages adolescents with substance use problems from seeking help. A mental health expert suggested that substance use should be decriminalized, though not legalized, to facilitate access to treatment for adolescents who use substances (see **Table 2, Quote 17)**.

### 3.3. Strategies to enhance the SBIRT program’s implementation

Guided by the SBIRT program vignette (See Study Box 2), participants shared their concerns and recommendations to enhance the program’s implementation. **Table 3** summarizes participant-identified strategies for enhancing the SBIRT program’s anticipated implementation outcomes guided by RE-AIM: Acceptability / Adoption and Reach. In terms of acceptability / adoption, most participants expressed concerns about the program being accepted by adolescents given its novelty and the current lack of treatment programs supporting those who use alcohol. Healthcare workers recommended conducting dissemination campaigns prior to launching the program in order to familiarize community members with its objectives and importance. Adolescents were concerned about stigma against those who use alcohol and / or live with HIV, potential breaches of confidentiality and mistreatment from healthcare workers. Community healthcare workers recommended making the alcohol screening mandatory to all adolescents, while adolescents recommended having peer adolescents provide counseling during screening (See Table 3). In terms of reach, recommendations included integrating the SBIRT program into existing community-based programs and contacting adolescents at alcohol hotspots (See Table 3).

**Table 3:** Recommended strategies for implementing an alcohol SBIRT program for adolescents, organized by selected RE-AIM implementation outcomes (Acceptability and adoption, Reach)

| Selected RE-AIM implementation outcomes | Participant-identified strategies to implement the SBIRT program | Selected exemplary quotes |
| --- | --- | --- |
| Acceptability and adoption | Disseminate information about the SBIRT program prior to launching it | *“I think that's a good program. So, I would urge the people are organizing this program to make sure that* |
|  | (e.g., social media, communities, schools)<br>• Make the program ‘mandatory’ to all adolescents living with HIV to reduce stigma against those who use alcohol<br>• Ask for informed consent prior to collecting urine sample<br>• Provide clear explanations and counseling both after and prior to collecting urine sample<br>• Protect confidentiality and trust by having specific healthcare workers delivering the SBIRT program<br>• Have peer adolescents provide counseling before and after screening, using empathetic, non-judgmental language | *if it's something that one has to do with youth or adolescence, they should also make sure that they should involve youth and adolescents in that program, because youth are free with their fellow youth. Mostly. Yes. Not all, but mostly they are free with their fellow youths”* (FGD, Adolescent)<br>*“-What do you think, should this program that might be introduced in HIV care be mandatory or voluntary to HIV adolescent patients or clients? - I think it should be mandatory, as they come for their routine services”* (FGD, Adolescent)<br>*“Adolescents need trust and assurance that the information they give you, you're going to keep it confidential, and that way they open up to you”* (FGD, Adolescent)<br>*“So the reason why we some of us don't hear to some counseling that being offered to us because most counselors are most people that do the counseling the peer educators, they are so judgmental. Okay, they make it seem as if they're, they said they are the spokesperson to Jesus Christ. That's how they make it seem”* (FGD, Adolescent) |
| **Reach** | • Deliver the SBIRT program through existing community-based organizations (e.g. churches)<br>• Expand the SBIRT program beyond HIV clinics to include preventative programs as a way to reduce HIV-related stigma and promote wider participation in the program (e.g. libraries)<br>• Reach adolescents within hotspots where they use alcohol or substances<br>• Integrate the SBIRT program into existing youth services (e.g. Youth Friendly Spaces)<br>• Design the SBIRT program in such a way to involve male adolescents, in addition to female adolescents | *“Personally, I think decentralizing the service would cater to a lot of adolescents. There's a previous study that was done with decentralized sexual reproductive health services from the facility, we brought them near to the community. So, they would have hubs, where adolescents would go and access health services. From my conclusion, I would say that the hubs drew more adolescents and young people as compared to the facility”* (FGD, Community healthcare worker)<br>*“We want this program to work, we also want a program for men, just like the way women have Dreams program, we also want men to be kept busy”* (FGD, Adolescents) |

## 4. Discussion

This qualitative study aimed to identify the multi-level contextual factors relevant to the implementation of a biomarker-based alcohol SBIRT program tailored to HIV-affected adolescents living in Lusaka, Zambia. We engaged a wide range of participants, including adolescents, caregivers, healthcare providers and policymakers, in the identification of implementation strategies for the future SBIRT program. Our analysis led to three key insights related to the implementation of the biomarker-based alcohol SBIRT program. First, the program was perceived as a potentially appropriate approach to identifying and treating adolescents with unhealthy alcohol use. Second, to ensure its acceptability and adoptability, the program’s implementation strategies should target relevant contextual factors of implementation, such as stigma and trust in healthcare providers. Third, to ensure that the program reaches at-need adolescents, it should capitalize on existing HIV care services as well as other existing alcohol and substance use prevention programs for adolescents. In addition, the program’s implementation strategies should be designed with an eye toward engaging both male and female adolescents from the start.

Our study suggests that a biomarker-augmented SBIRT program may be an appropriate approach to start addressing unhealthy alcohol use among adolescents in Lusaka, Zambia, a public health concern for participants. In line with previous nationally representative surveys showing that the prevalence of unhealthy alcohol use among HIV-affected young adults is 21.6% [33], our qualitative assessment suggested that there is an urgent need to implement programs focused on early identification and treatment of unhealthy alcohol use for adolescents. Our study also highlighted that the wider population of adolescents beyond those affected by HIV are also in need, particularly those exposed to structural determinants of alcohol use, including poverty, violence, and other traumatic childhood experiences. The biomarker-based SBIRT program would be the first one of its kind in a context where previous programs have failed or focused on the two extremes of the alcohol use spectrum: primary prevention or in-hospital treatment for alcohol intoxication. As the first SBIRT program for HIV-affected adolescents, it should be carefully designed with a focus on addressing key contextual factors that would facilitate its successful implementation in Lusaka, Zambia.

Our assessment of implementation context highlighted that stigma is a widespread factor that would need to be addressed for the SBIRT program to be acceptable and adoptable among adolescents. We identified at least two sources of stigma related to the program implementation. First, in line with previous studies [35], we found strong and widespread stigma against adolescents who use alcohol or other substances in Zambia, who are often seen dangerous people that should be kept at distance. Such perceptions may be fueled by the lack of treatment for adolescents with unhealthy alcohol use, which often prevents adolescents from reducing alcohol use, keeping them at risk of developing an alcohol use disorder. Second, we identified HIV-related stigma, including stigma against those with HIV infection, antiretroviral use, and use of HIV services. Previous studies have documented multiple sources of HIV-related stigma experienced by adolescents in Zambia, including internalized, community-and health system-level stigma [36, 37]. HIV-related stigma seems to negatively impact adolescents’ access to HIV services and antiretroviral use. Moreover, studies have suggested that stigma may be compounded [38]. For instance, a previous study conducted in South Africa, indicated that people who experience internalized stigma for both HIV and unhealthy alcohol were particularly affected by negative mental health outcomes and functional impairment [39]. Along these lines, our study indicated that, if unaddressed, the fear of being labeled ‘alcoholics’ may deter adolescents from engaging in the SBIRT program and limit their participation in HIV care services, which aligns with previous qualitative studies [40]. Alternatively, the fear of being labeled ‘HIV-positive’ could limit adolescents’ participation in the SBIRT program. Participant-identified strategies for addressing stigma related to program participation included implementing a communication campaign to describe the objectives of the program and the need for addressing alcohol use among adolescents prior to launching the program; making the alcohol screening phase universal, similar to screening for HIV and sexual transmitted infections, to avoid labeling adolescents who use alcohol; and integrating the screening phase into routine healthcare processes beyond HIV care as a way to avoid identifying adolescents living with HIV.

Adolescents’ trust in healthcare providers was identified as a relevant contextual factor that may influence the program’s acceptability. In line with previous research, our study suggested that providers may contribute to reducing or further replicating stigma against alcohol use and other health conditions [40, 41]. The providers’ communication style, attitudes, and behaviors prior, during and after the screening phase of the SBIRT program may influence the adolescents’ initial willingness to participate, as well as their subsequent engagement in the program. Providers’ behavior during the initial screening of the SBIRT program seemed particularly relevant. Participant-identified strategies related to trust in healthcare providers included building rapport by carefully explaining why the alcohol screening is important and why the urine sample is needed; utilizing compassionate attitudes and language, while avoiding authoritative, paternalistic, or judgmental behaviors; and respecting the adolescents’ privacy and confidentiality during the screening and counseling phases. The implementation infrastructure, particularly the availability of private spaces within clinics to conduct the screening and brief counseling phases of the SBIRT program, is also essential to ensure the program’s acceptability among adolescents.

Our study suggested that the degree of shared identities and experiences between adolescents and healthcare providers may also influence the program’s acceptability from the perspectives of adolescents. One of the most salient implementation strategies emphasized by adolescents consisted of having youth peer counselors deliver the screening and counseling phases of the SBIRT program, with counseling before and after screening. This was different to our original vignette (See Study Box 2), in which a nurse provided counseling just after the screening phase. Peer support has been increasingly featured by WHO as part of adolescent-friendly spaces as an equitable, acceptable, and appropriate approach to addressing health outcomes such as engagement in HIV treatment [42]. Studies have suggested that peer-led approaches may be particularly acceptable during adolescence as they may protect from the effects of stigma, influence behavior, instill hope, and share coping strategies based on common experiences and backgrounds [43]. Previous studies conducted in Zambia and other countries in Southern Africa have shown that peer-led support may be an effective and acceptable approach to addressing HIV-related health outcomes among adolescents [43, 44]. Future studies should focus on defining the methods for selecting and training peers and evaluating the effectiveness of peer-led screening and brief intervention programs for unhealthy alcohol use among adolescents. Studies should also assess the feasibility and added benefits of providing counseling both before and after screening, as suggested by participants, compared to after screening only.

While our study found a scarcity of treatment programs for adolescents with unhealthy alcohol use, our results suggest that there is a robust healthcare delivery infrastructure that could serve as the foundation to implement a biomarker-augmented SBIRT program. Our study confirmed that available HIV care services are reaching HIV-affected adolescents through clinic- and community-based treatment and counseling programs. Future studies should focus on mapping out the specific healthcare processes in which the SBIRT program may be integrated into. Our study also identified limitations in the existing implementation infrastructure. Results suggest that the adolescent population in need of alcohol treatment programs goes beyond the HIV-affected adolescent population. Integrating the SBIRT program through HIV care services only may leave out a wide proportion of adolescents, particularly those affected by structural determinants of alcohol use who often lack access to healthcare services. To address this issue, participant-identified implementation strategies included integrating the SBIRT program into broader services aimed at addressing social determinants of health, such as poverty, through vocational training; and conducting alcohol screening and brief interventions at community-level hotspots where adolescents commonly use alcohol, such as schools or vacant lots. We also found that most existing preventative programs for adolescents seem to prioritize reaching girls over boys. Our results point to gender-based differences in the motivations for and behaviors of alcohol use among adolescents, with girls often using alcohol during sex as they believed it enhances boys’ satisfaction. Participants emphasized that the program should aim to reach boys and girls through tailored implementation strategies developed during the design phase of the program.

Results from this study should be interpreted considering three main limitations. First, the parent qualitative study was designed to describe the context of adolescent alcohol and substance use and the participants’ perceptions of a novel biomarker-based alcohol SBIRT program. This secondary analysis applied implementation science themes to previously collected qualitative data. Second, the number of policymakers (n=7) was small due to the limited availability of policymakers who are knowledgeable about alcohol use in Zambia. Third, results from this study, obtained in Lusaka, Zambia, cannot be directly applied to other settings. However, the knowledge generated through this study sheds light on the need for and potential implementation strategies of other alcohol SBIRT programs in Zambia. While our study focused on an alcohol SBIRT program, insights obtained through this study may also help to design future intervention programs for other unhealthy substance use among adolescents in Zambia. Future studies should also explore how to approach discrepancies between biomarker-based alcohol screening and self-report alcohol screening.

## 5. Conclusions

An alcohol biomarker-augmented SBIRT program may be an appropriate, acceptable, and adoptable approach to addressing unhealthy alcohol use among adolescents in Lusaka, Zambia. Implementation strategies of the program should address contextual factors. Our study showed that the implementation of the SBIRT program would touch upon aspects of compounded stigma related to HIV and alcohol use among adolescents. Using an alcohol biomarker during the screening phase would help to address under-reporting but would require the careful design of strategies to ensure adolescents’ trust and engagement in the program. At follow-up appointments, the alcohol biomarker could motivate behavioral change, reducing unhealthy alcohol use. Delivering youth peer-led counseling prior to the screening and during the brief intervention phases of the SBIRT program is a strategy that may promote the program’s acceptability and adoptability. Integrating the program into existing HIV care services and other preventative programs for adolescents, as well as designing strategies that engage both males and adolescents from the start, may help to enhance the program’s reach.

## Data Availability

Data will be made available upon reasonable request.

## 6. Acknowledgements

We wish to thank the community workers who contributed to this study and the study participants that shared their time, experiences and recommendations with our research team.

## 9. Authors’ contributions statement

**Paniagua-Avila**: Conceptualization, Methodology, Formal analysis, Writing – original draft, Writing – review and editing; **Kanguya**: Formal analysis, Investigation, Project administration, Supervision, Writing – review and editing; **Mwamba**: Formal analysis, Supervision, Writing – review and editing; **Hahn:** Conceptualization, Writing – review and editing; **Latkin**: Conceptualization, Writing – review and editing; **Chander**: Writing – review and editing; **Martins**: Writing – review and editing; **Munthali**: Writing – review and editing; **McDonnell**: Writing – review and editing; **Sharma**: Conceptualization, Methodology, Resources, Project administration, Supervision, Writing – original draft, Writing – review and editing; **Kane**: Conceptualization, Methodology, Resources, Writing – original draft, Writing – review and editing, Supervision, Funding acquisition

